# A bibliometrics analysis about male prepuce with Machine Learning

**DOI:** 10.1101/2023.10.31.23297777

**Authors:** Bibo Lan, Mengling Nian, Qian Liu, Xinyi Cai

## Abstract

Limited research has been conducted on the male prepuce. Despite its ana-tomical complexity and role as an erogenous structure that interacts with adjacent penile structures, there is a dearth of information on this topic. To address this gap, this study employs bibliometric analysis to identify primary research areas and trends related to the male prepuce over the past two decades. A systematic search was performed on the Web of Science core collection to identify publications related to the male prepuce that were published between 2003 and 2022. Following this, bibliometric analysis and visualization were conducted utilizing the bibliometrix R package, the text2vec R package, Citespace, and VOSviewer software. This study presents a novel compilation and review of the authors, country, institutions, journals, and keywords pertaining to the male prepuce in the literature. The scarcity of adult and child prepuce pathologies, likely due to the prevalence of circumcision in many societies, may have hindered previous recognition of the academic significance of this anatomical feature. Through bibliometric analysis, this study investigates research trends and highlights current areas of interest, as well as identifying the most prolific authors, countries, institutions, and journals in this field.

## Introduction

Few independent studies have been conducted on the male prepuce, which is often excised during circumcision in various cultures^1^.

The male prepuce, or foreskin, serves as a junctional mucocutaneous tissue that plays an important role in the external aspect of male genitalia. Despite being frequently overlooked, this function of the prepuce is significant^2^.

The double layer of epithelium in the penis protects the glans and urinary meatus from abrasion, friction, and damage, making it a vital protector^1^. Additionally, the organ has specialized immune cells, lymphatic vessels, and exocrine glands that produce lytic material, contributing to its immunological function^3^. The frenulum of the male prepuce contains thousands of fine-touch receptors and pheromone-producing apocrine glands that enhance the penis’s erogenous function^4^. During erection, approximately half of the smooth muscle sheath of the male prepuce elongates, accommodating the entire penis^5^.

The inadequate development of the male prepucehas been found to be correlated with additional penile abnormalities, including hypospadias, chordee, and epispadias^6^. In boys with hypospadias, during weeks 8-14 of pregnancy, the opening of the urethra does not form near the penis’ tip. Abnormal openings can occur anywhere under the penis to the scrotum^7^. Chordee is a pathological condition characterized by a downward or upward curvature of the penile head at the junction with its shaft. During erection, the curvature of the penis is prominently noticeable, however, even in a flaccid state, resistance to straightening can also be observed. Epispadias is a rare form of urogenital malformation in which the urethral tube fails to tabularize on the dorsal side^8^.

While the male prepuce is of great anatomical and physiological importance, limited academic publications have tried to understand the research pattern related to the male prepuce systematically. An important aspect of bibliometrics is that it provides a means for analyzing large amounts of scientific information in a rigorous manner. Until now, no one has collected, reviewed, or analyzed the authors, countries, institutions, journals, and keywords associated with the male prepuce.

## Materials and methods

### Search strategy

The data for our study on the male prepuce was obtained from the Web of Science Core Collection database. We searched for publications related to the topic that were published between 2003 and 2022 as of May 31, 2023. Our search strategy included the terms ‘foreskin’ or ‘male prepuce’, resulting in a total of 4453 studies. From these, we collected 2627 literature data that included articles and reviews, which were downloaded from WOS core with full records and cited references (see Figure 1). We then performed a visual analysis of the data.

**Figure 1.**
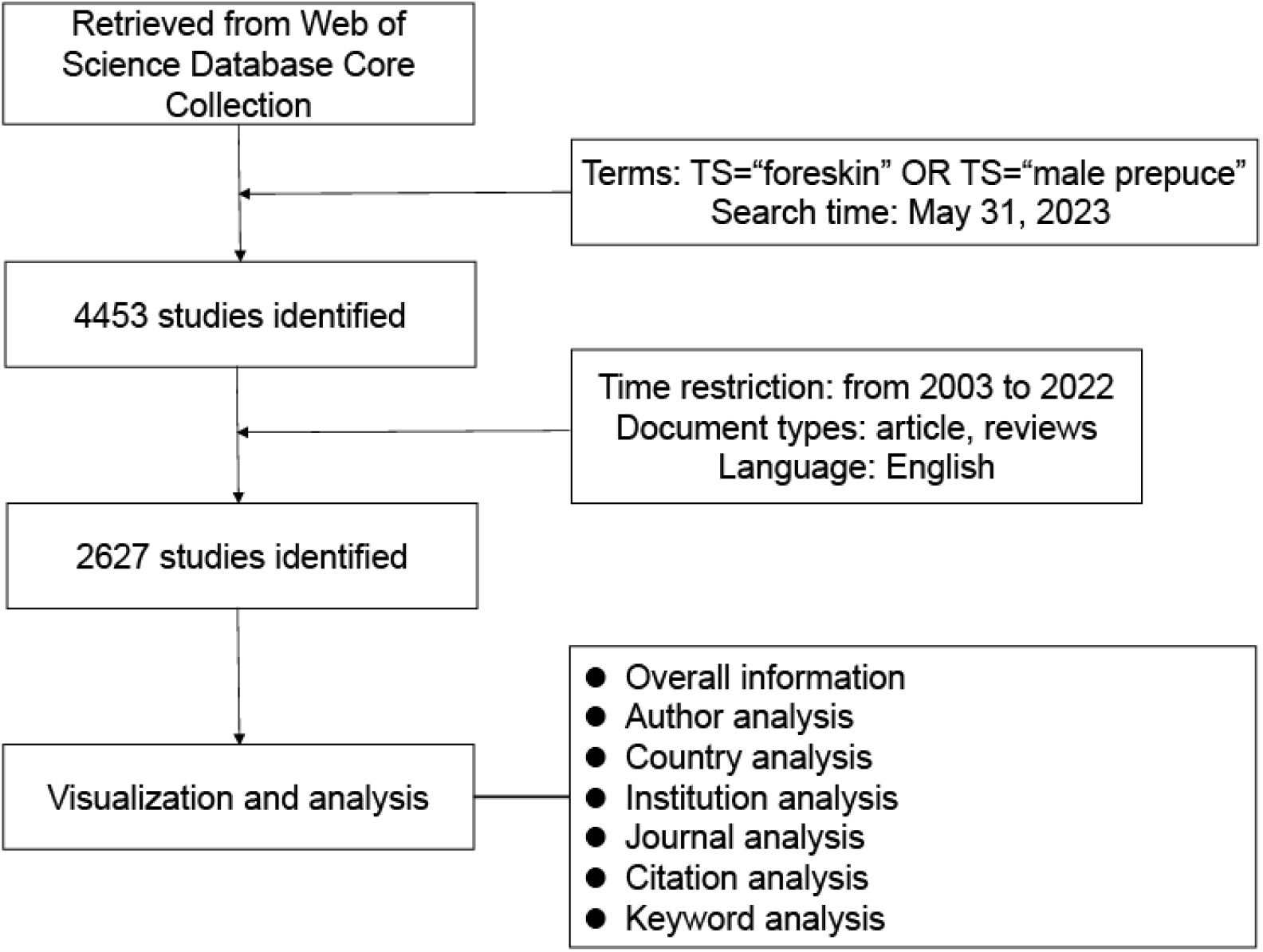
Flow chart in male prepuce

### Visualization and analysis

In this study, we analyzed the data using bibliometrix v4.0.0 R package, online bibliometric analysis platform (https://bibliometric.com), text2vec R package, Citespace (version 6.1.6), and VOSviewer (version 1.6.19). The bibliometrix v4.0.0 R package and online bibliometric analysis platform (https://bibliometric.com) provided a comprehensive analysis for the analysis of the annual publication, authors, country, institutions, and journals. And it was used to obtain the thematic mapping and milestone literature concerning the publications of the field of the male prepuce research.We used Citespaces software which is a bibliometric visualization software to plot a dual-map overlay of journals. Vosviewer software was employed to visulize network based on the relationship between country. For abstract analysis, we used the text2vec R package to conduct Linear Discriminant Analysis (LDA). We extracted each word from the summary, built a term-co-occurrence matrix, and constructed the LDA topic model.

## Results

### The analysis of annual publication and citation

Over the past two decades, there has been a growing interest in the field of the male prepuce study, as evidenced by the annual output of publications. Notably, there was an upward trend in the number of articles published, with fewer than 100 articles published from 2005 to 2007 and more than 100 published from 2010 to 2022. Additionally, the highest average article production occurred in 2022 (182), as shown in Figure 2.

**Figure 2.**
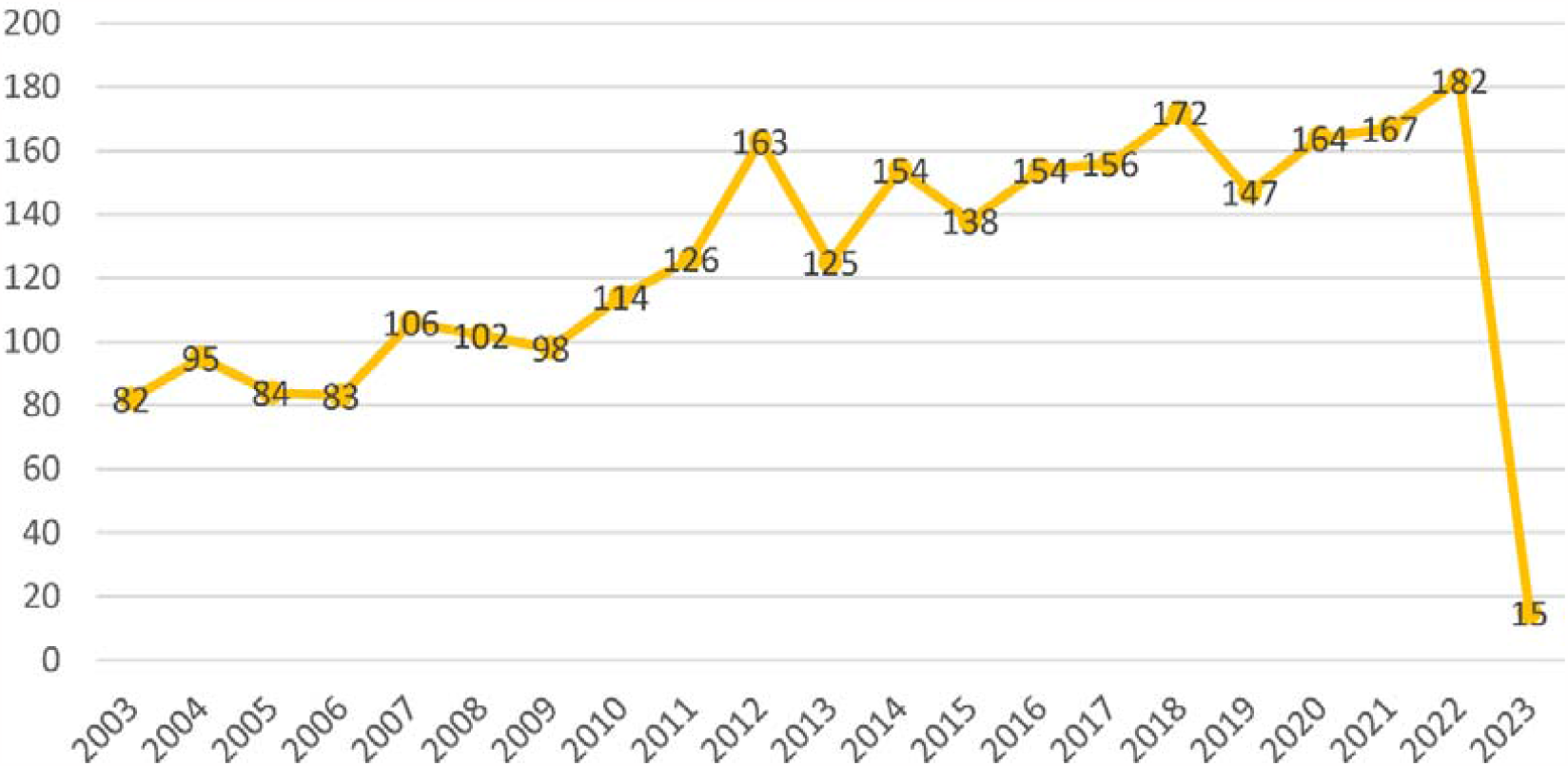
Annual production from 2003 to 2022 in male prepuce

### Author analysis

Figure 3 displays the top 20 authors with the highest productivity over time. The size of the dot corresponds to the number of articles, while the color represents the number of citations. Wang X exhibited a consistent trend and had the most articles (29), followed by ZHANG Y (28) and WANG Y (27). Wang X also had the highest total number of citations, with CUBILLA AL coming in second. Notably, 12 of the top 20 authors are from China. WANG X also had the highest h index and g index. The detail description is show in Supplement Table 1.

**Figure 3.**
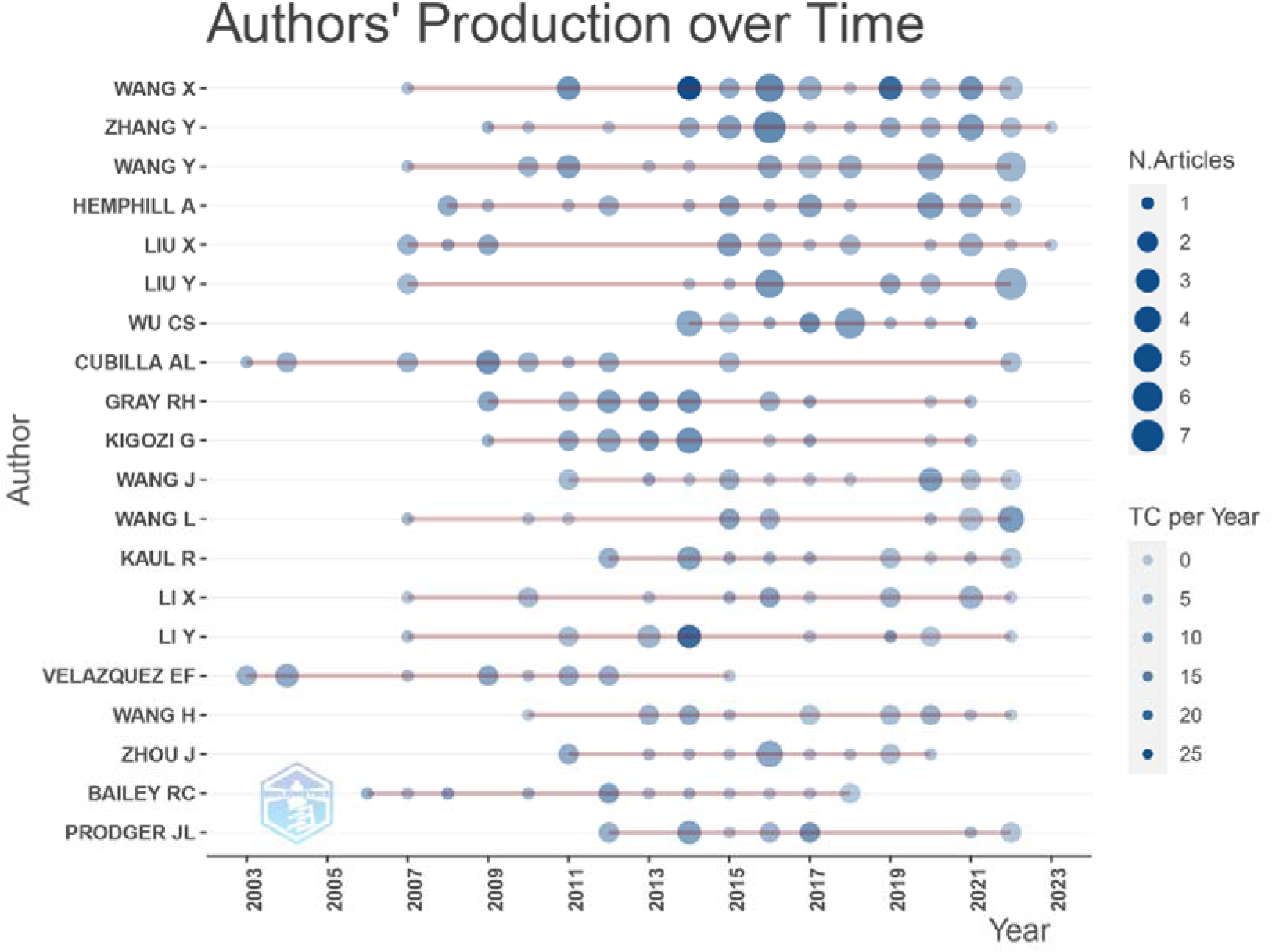
Authors’ production over time from 2003 to 2022 in male prepuce

### Country analysis

In this study, Figure 4 displays the number of publications by country, with circle size representing the quantity and line distance indicating collaboration intensity. As shown in Figure 4 (A) The United States had the most publications, followed by China and Germany. The top five countries were the United States (845), China (420), Germany (183), the UK (169), and Iran (122). Besides, the top seven countries had published more than 100 articles. Figure 4 (B) shows that among the top 20 countries, developed countries such as the USA, Germany, and Japan were the first to conduct research in this area. However, in recent years, China, Italy, Spain, and other countries have become more focused on this field.

**Figure 4.**
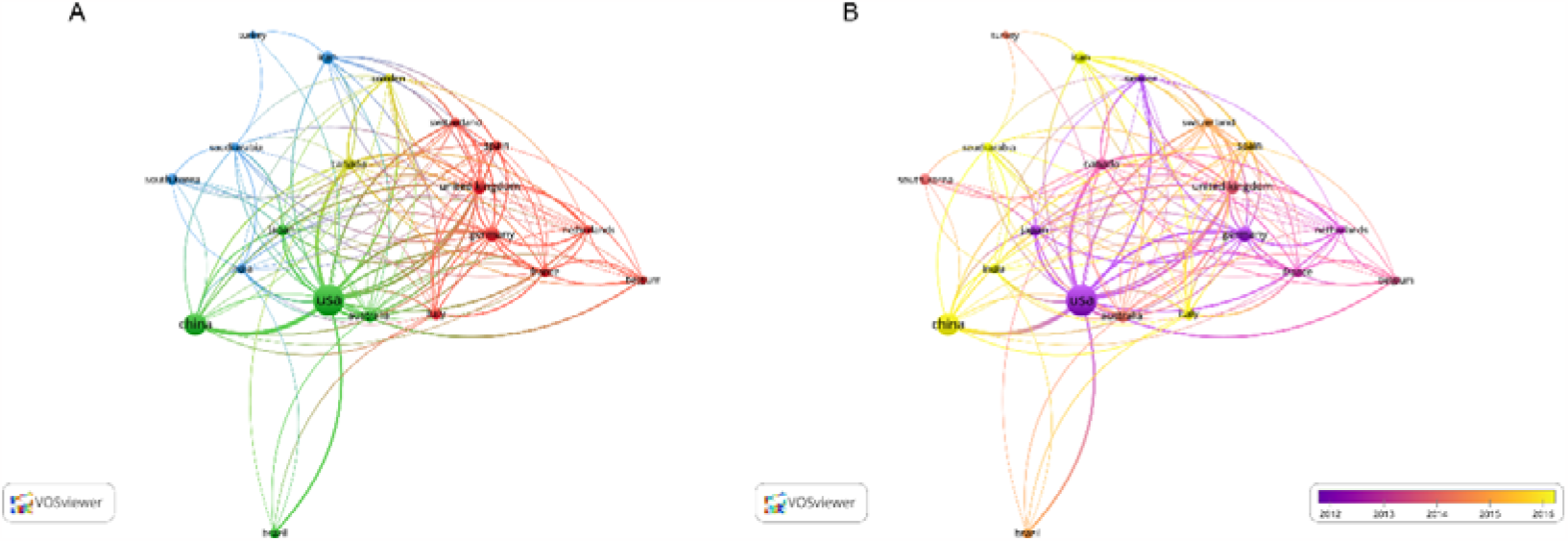
Co-authorship analysis in countries (A) The publication in top 20 countries (B) The publication in top 20 countries over time

### Institution analysis

Figure 5 shows the top 20 institutions in terms of publication number. The University of California, San Francisco had the highest number of publications (112), followed by Johns Hopkins University (105), University of Washington (93), University of Bern (82), and Shanghai Jiao Tong University (75).

**Figure 5.**
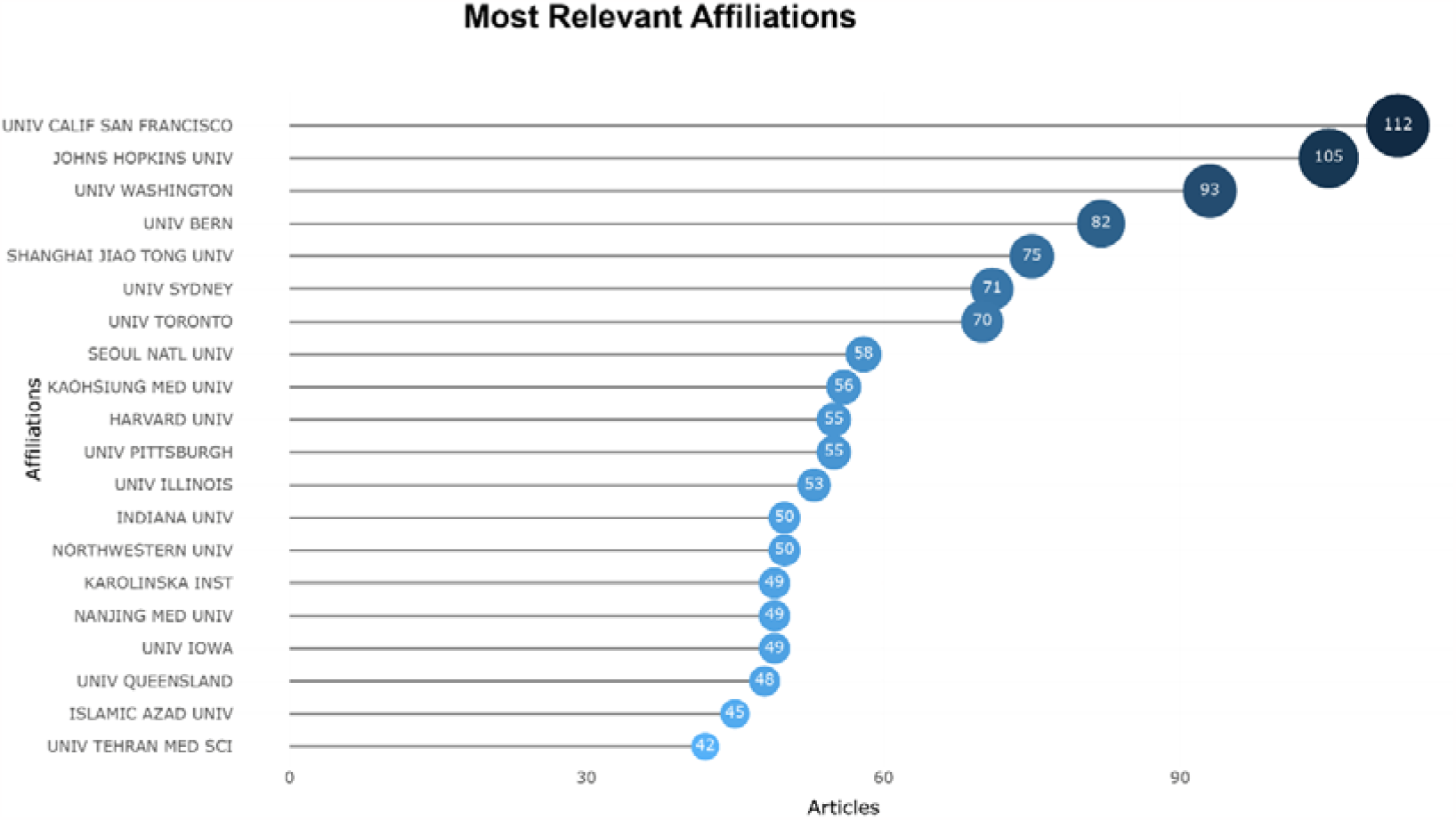
The documents in top 20 institutions

### Journal analysis

Figure 6 shows the top 20 journals in terms of publication number. PlOS One had the highest number of publications, followed by JOURNAL OF VIROLOGY, JOURNAL OF PEDIATRIC UROLOGY, JOURNAL OF UROLOGY, and UROLOGY. Most of the studies related to male prepuce are published in general interest journals.

**Figure 6.**
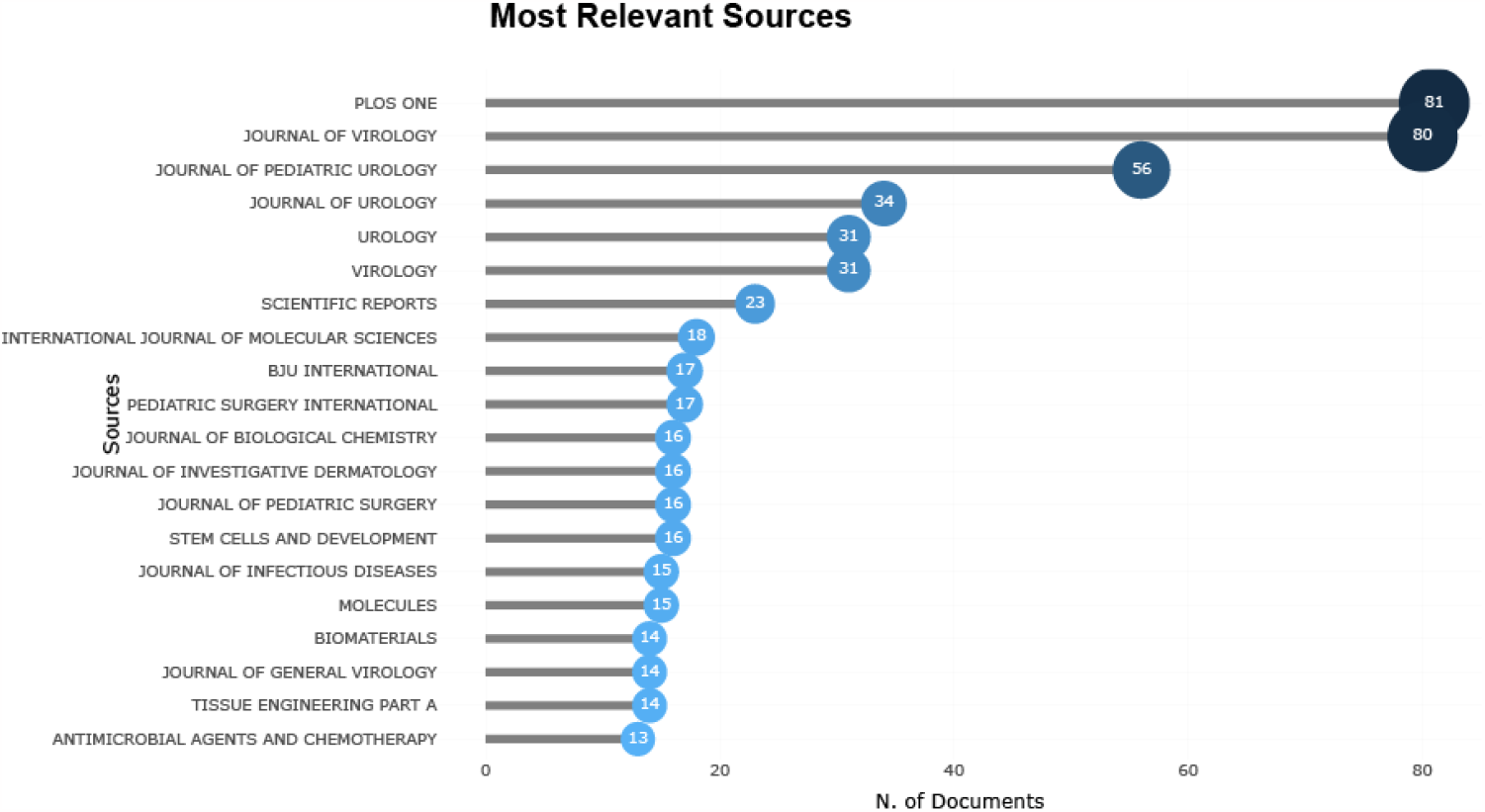
The numbers of documents in top 20 journals.

In our study, we employed a dual-map to visually present the primary citation paths, which effectively demonstrated the distribution of topics in the journals. The map exhibited the citing journals on the left side, cited sources on the right, and utilized color stripes in the middle to indicate the citation paths. Figure 7 displays four main citation paths, two of which are green and the other two yellow. The green paths revealed that studies published in MEDICINE, MEDICAL, CLINICAL journals were commonly cited in MOLECULAR, BIOLOGY, GENETICS and HEALTH, NURSING, MEDICINE journals. Additionally, the yellow paths revealed that studies published in MOLECULAR, BIOLOGY, IMMUNOLOGY were frequently cited in MOLECULAR, BIOLOGY, GENETICS journals.

**Figure 7.**
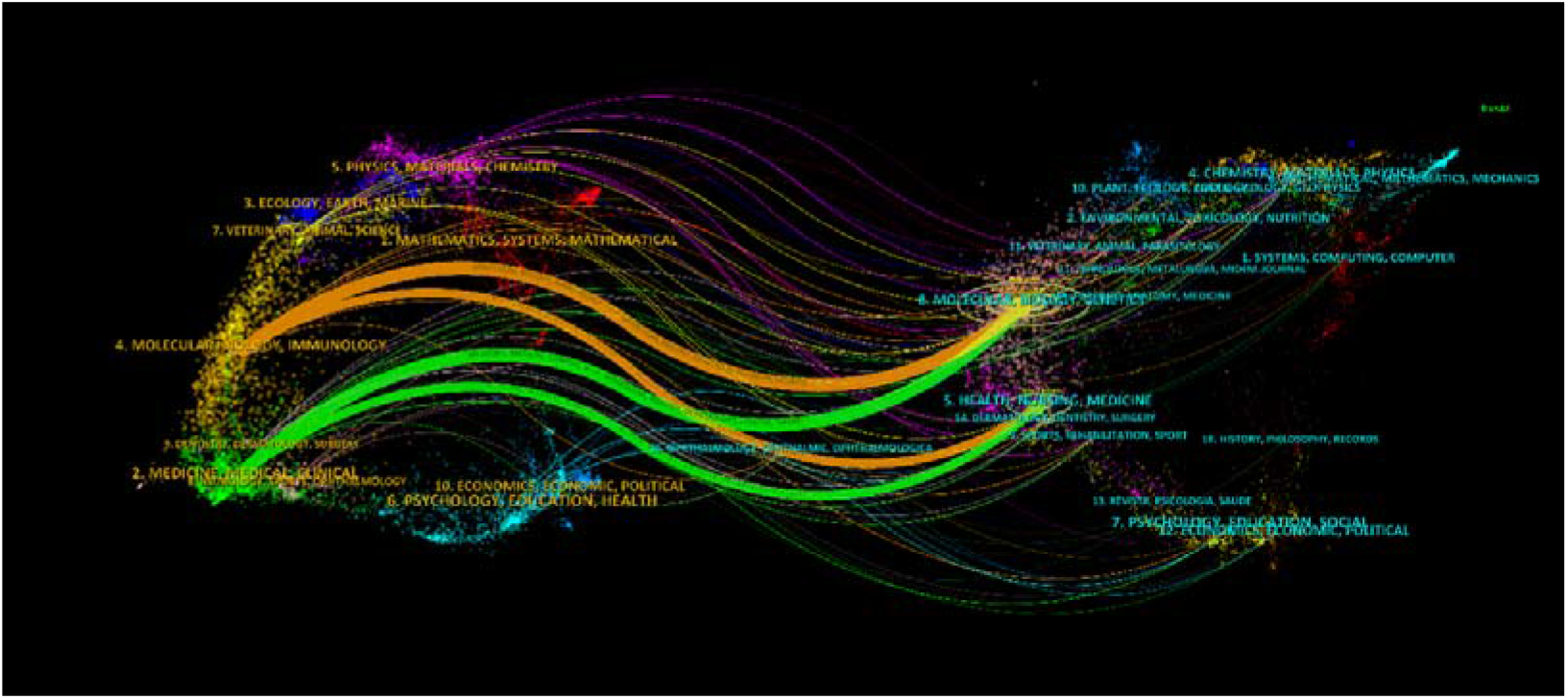
A dual-map between citing journals and citing journals from 2003 to 2022 in male prepuce

### Citation analysis

Supplement Table 2 displays the top 10 citations related to male prepuce research, with A CULTURE SYSTEM USING HUMAN FORESKIN FIBROBLASTS AS FEEDER CELLS ALLOWS PRODUCTION OF HUMAN EMBRYONIC STEM CELLS having the highest global citation score (GCS). GCS is defined as the number of citations in the Web of Science database.

### Abstract analysis

Based on the intertopic distance map generated by LDA for a three-topic model, it can be observed that the three topics remained distinct from each other as they did not overlap, as depicted in Figure 8A, B, C. The study found that the represented topics in the research were proportionally distributed, with 40.2% for abstract Topic 1 (Figure 8A), 30.1% for abstract Topic 2 (Figure 8B), and 29.7% for abstract Topic 3 (Figure 8C). The most representative terms in Topic 1 were related to gene expression patterns of the foreskin, including ‘cells’, ‘human’, ‘expression’, and ‘gene’. Topic 2 focused on the surgical removal of the foreskin (circumcision) and included terms such as ‘patients’, ‘circumcision’, and ‘treatment’. Topic 3 focused on the bioengineering application of foreskin and included terms such as ‘differentiation’, ‘epidermal’, and ‘based’.

**Figure 8.**
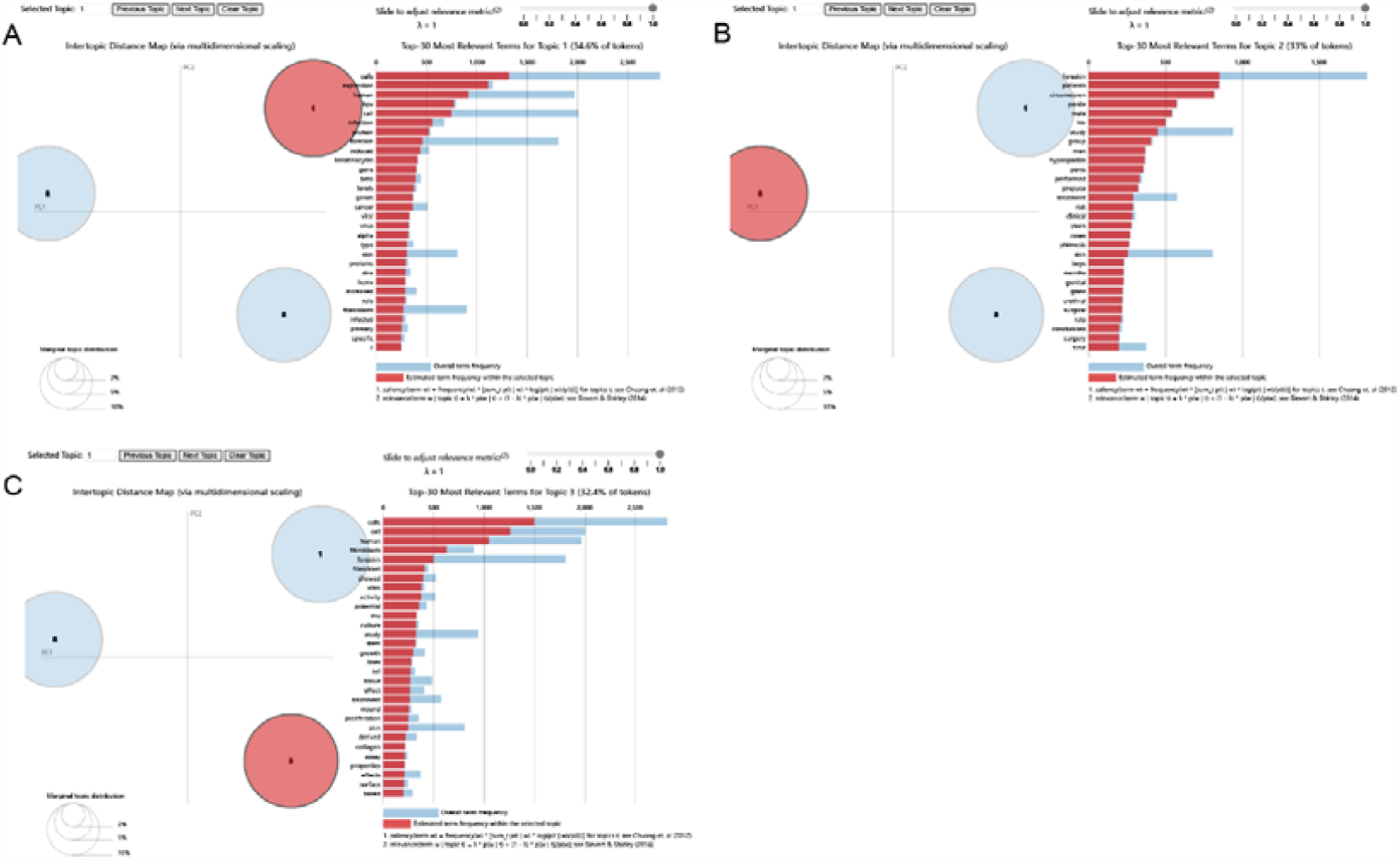
This figure depicts the intertopic distance maps generated by LDA analysis for three different topics related to foreskin. Figure 8(A) shows the gene expression pattern of the foreskin, with the size of the red circle representing the proportion of Topic 1 information in the overall texts and each red bar presenting a term frequency in Topic 1. Figure 8(B) shows the intertopic distance map for the surgical removal of the foreskin (circumcision), with the size of the red circle representing the proportion of Topic 2 information in the overall texts and each red bar presenting a term frequency in Topic 2. Figure 8(C) shows the intertopic distance map for the bioengineering application of foreskin, with the size of the red circle representing the proportion of Topic 3 information in the overall texts and each red bar presenting a term frequency in Topic 3.

### Keyword analysis

The thematic map, illustrated in Figure 9, was created using author keywords and organized into four distinct themes: niche (located in the left top quadrant), motor (located in the top right quadrant), emerging or declining (located in the left bottom quadrant), and basic themes (located in the right bottom quadrant). The motor themes, which are well-established areas of research, are plotted in the top right quadrant and include the following topics: fibroblasts, apoptosis, and keratinocytes. Moreover, there are basic themes (circumcision, foreskin, hypospadias) in most scholarly production of the male prepuce research. Niche themes are represented in cluster: tissue engineering, chitosan, electrospinning. In emerging or declining themes, Cluster contains theme concerning toxopiasmagohdi.

**Figure 9.**
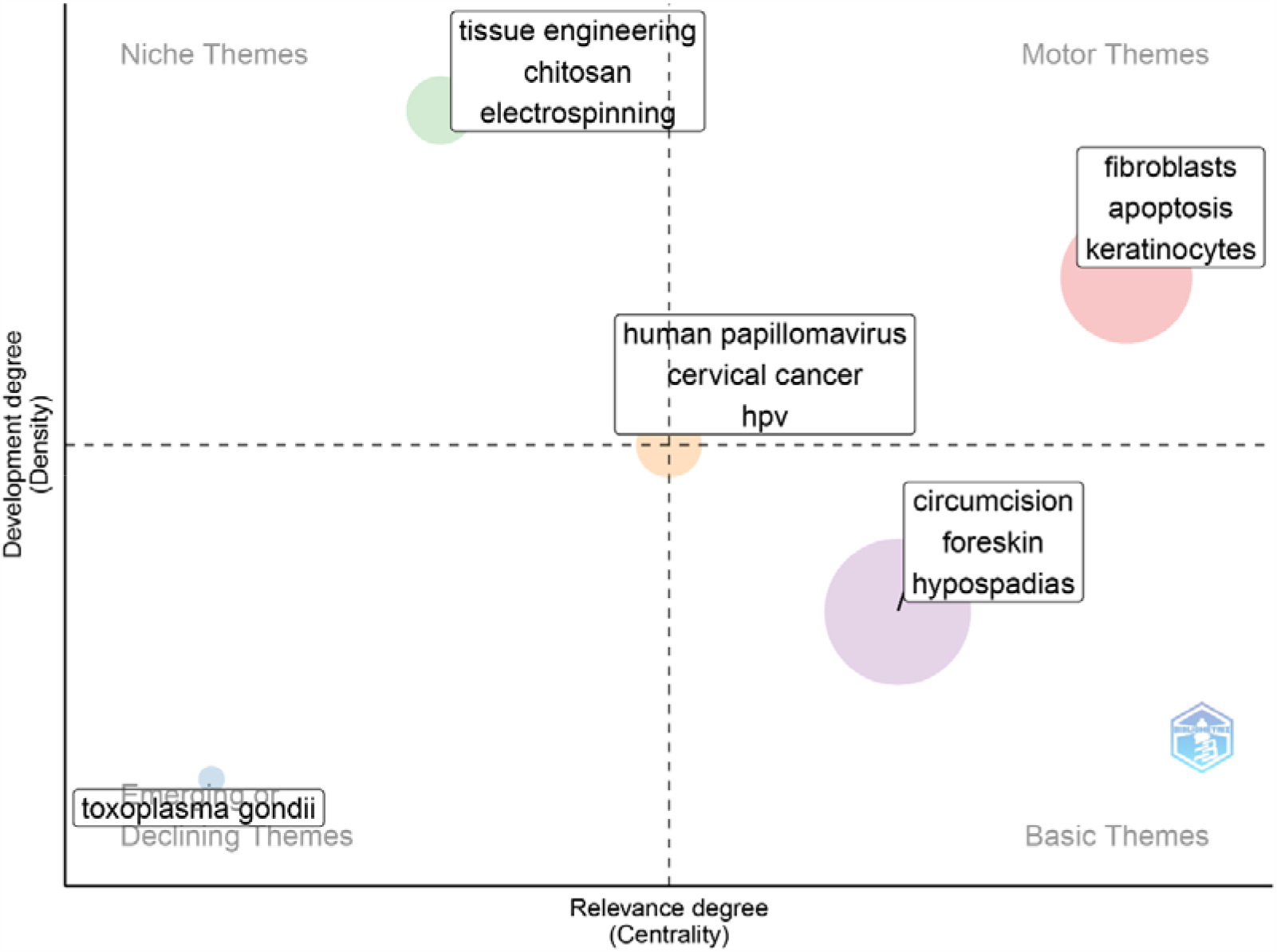
Visualization of thematic mapping

## Discussion

This study presents a bibliometric analysis of male prepuce publications over the past 20 years, investigating global productivity and research trends. The results reveal a strong interest in this field, with an increasing trend in annual publications. The United States is the leading contributor to foreskin research, likely due to strong support from funding agencies. Wang X is the most prolific author with a consistent publication trend. PLOS ONE is the most productive journal, and The University of California, San Francisco has the highest number of published papers among all institutions. Our research utilized a dual-map to present the primary citation pathways, showcasing the distribution of topics among various journals. Our findings indicated that publications in MEDICINE, MEDICAL, and CLINICAL journals were frequently referenced in MOLECULAR, BIOLOGY, GENETICS, as well as HEALTH, NURSING, and MEDICINE journals. Additionally, studies published in MOLECULAR, BIOLOGY, and IMMUNOLOGY journals were commonly cited in MOLECULAR, BIOLOGY, and GENETICS journals.

Thematic mapping research has highlighted the core position of Human papillomavirus (HPV), cervical cancer, and HPV in the field, with related research spanning ancient times to the present. In 2008, a study revealed that the prevalence of HPV infection in the glans was significantly higher in uncircumcised men than in circumcised men, even after adjusting for demographic characteristics and sexual history. Additionally, uncircumcised men had a higher risk of oncogenic HPV infection and infection with multiple HPV types^9^. HPV infection is strongly associated with penile cancer, lack of neonatal circumcision, and phimosis^10^.

However, male circumcision can reduce the risk of oncogenic HPV genotypes, cervical cancer, T. vaginalis, bacterial vaginosis, and possibly genital ulcer disease in women^11^. In motor themes, research themes include cluster: fibroblasts, apoptosis, keratinocytes. Niche themes are represented in cluster: tissue engineering, chitosan, electrospinning. These two fields of male prepue (foreskin) research have reached a relatively mature stage. Fang Fang and colleagues have proposed a standardized and repeatable method for isolating fibroblasts from human foreskin tissues and identifying their biological characteristics^12^. Previous studies have also indicated that apoptosis plays a crucial role in the development of male genitalia^13^. Furthermore, researchers have successfully used foreskin-isolated keratinocytes for extemporaneous autologous pediatric skin grafts^14^. Hypospadias, a common birth defect in boys, has prompted the exploration of various biomaterials for urethral repair, including synthetic and natural polymers^15^. The research on the male prepuce typically revolves around themes such as circumcision, foreskin, and hypospadias.

These topics are considered as research hotspots and potential areas for future development. In emerging or declining themes, Cluster contains theme concerning toxopiasmagohdi. The incidence and prevalence of Toxoplasma infection have significantly decreased over the past 30 years, resulting in a gradual decline in research on Toxoplasma gondii^16^.

The top three most cited publications on the male prepuce discuss the application of fibroblasts. Human embryonic stem (hES) cells show promise for use in research areas such as human developmental biology and cell-based therapies^17-19^. These cells are traditionally cultured on mouse embryonic fibroblast (MEF) feeder layers, allowing continuous growth in an undifferentiated stage. However, for use in human therapy, an animal-free culture system must be utilized to prevent exposure to mouse retroviruses. To ensure successful human embryonic stem cell (hESC) transplantation and prevent immune rejection and zoonotic transmission, it is crucial to eliminate animal-derived constituents, nonhuman sera, and animal feeder cells from hESC culture. In order to reduce the use of animal products, various human feeder cell types have been evaluated for hESC culture, including fetal muscle and skin cells, as well as adult fallopian tube epithelial and muscle cells. Human foreskin fibroblasts (hFFs) have proven successful in maintaining the derivation and undifferentiated growth of hESCs. One major advantage of foreskin feeders is their ability to be cultured continuously for over 42 passages. This allows for thorough analysis of foreign agents and genetic modification, such as antibiotic resistance. Additionally, it reduces the workload involved in continuously preparing new feeder lines.

The fourth, fifth, seventh, eighth, ninth most cited publications discussed HIV-1 target cells in the male prepuce^20-24^. Several epidemiological investigations have established substantial correlations between non-circumcision and the acquisition of HIV-1 in males. The study analyzed foreskin specimens from 20 males with a history of sexually transmitted infections and 19 males with no such history, in order to identify HIV-1 target cells. Many Langerhans cells were located in the epithelium, while the majority of CD4+ T cells and macrophages were situated in the submucosa.

A study on the male prepuce found that the fine-touch pressure thresholds in the adult penis were significantly lower in circumcised men than in uncircumcised men, after controlling for age, location of measurement, type of underwear worn, and ethnicity^25^. This indicates that the sensitivity of the glans of the circumcised penis is comparatively lower than that of the uncircumcised penis. Additionally, the most sensitive region of the uncircumcised penis is the transitional area between the external and internal prepuce, surpassing the most sensitive region of the circumcised penis. Therefore, circumcision results in the removal of the most sensitive parts of the penis.

The tenth most cited publications discussed the male circumcision^26^. Based on an assessment of existing evidence, it can be concluded that the advantages of male circumcision for newborns surpass the potential drawbacks, and therefore, families who opt for this procedure should have access to it. Randomized controlled trials have shown that male circumcision is associated with a lower prevalence of human papillomavirus (HPV) infection, herpes simplex virus type 2 (HSV-2) transmission, and bacterial vaginosis (BV) in female partners^27-29^. However, the evidence supporting male circumcision as a protective measure against syphilis is not as strong, and there is no connection between male circumcision and decreased risk of gonorrhea or chlamydia^30^.

In the field of LDA analysis, over 40% of studies have centered on examining gene expression patterns of the foreskin, while approximately 30% have focused on the surgical removal of the foreskin. The remaining studies have explored the bioengineering applications of foreskin. Interestingly, nine out of the top ten most cited articles involved foreskin-related cells, while the tenth publication discussed the significance of circumcision. Bioengineering has also been shown to play a crucial role in addressing the disease of hypospadias in boys.

### Limitation

This study only included articles and reviews from the WOSCC database, which may limit its representation of all current research on male prepuce. The research primarily focused on journals, with less attention given to other forms of scientific knowledge dissemination such as books, working papers, and reports. Therefore, some important emerging studies may have been overlooked.

## Conclusion

This bibliometric study presents a novel compilation and review of the authors, country, institutions, journals, and keywords related to the male prepuce, which has not been previously documented in the literature. The prevalence of circumcision in various societies may have hindered the recognition of the academic significance of the male prepuce, given the infrequency of adult and child prepuce pathologies. The study trends in male prepuce research were analyzed, and current research hotspots were identified.

## Data Availability

All data produced in the present study are available upon reasonable request to the authors

## Author’s Contribution

BB Lan: Manuscript writing

Mengling Nian: Data analysis

Qian Liu: Manuscript editing

YW Yue: Project development

## Conflicts of Interest

The authors declare no conflict of interest.

## Research involving Human Participants and/or Animals

Not applicable.

## Informed Consent Statement

Not applicable.

## Notes

### Competing Interest Statement

The authors have declared no competing interest.

### Funding Statement

This study did not receive any funding

