## Supplement Table for "A bibliometrics analysis about male prepuce with Machine Learning"

Supplement Table 1 The detailed of Authors’ production over time from 2003 to 2022 in male prepuce

|  | Element | h_index | g_index | m_index | TC | NP | PY_start |
| --- | --- | --- | --- | --- | --- | --- | --- |
| 1 | BAILEY RC | 8 | 13 | 0.444444 | 447 | 13 | 2006 |
| 2 | CUBILLA AL | 14 | 17 | 0.666667 | 613 | 17 | 2003 |
| 3 | GRAY RH | 13 | 17 | 0.866667 | 567 | 17 | 2009 |
| 4 | HEMPHILL A | 13 | 21 | 0.8125 | 459 | 23 | 2008 |
| 5 | KAUL R | 8 | 14 | 0.666667 | 268 | 14 | 2012 |
| 6 | KIGOZI G | 12 | 16 | 0.8 | 520 | 16 | 2009 |
| 7 | LI X | 8 | 14 | 0.470588 | 212 | 14 | 2007 |
| 8 | LI Y | 8 | 14 | 0.470588 | 366 | 14 | 2007 |
| 9 | LIU X | 11 | 20 | 0.647059 | 447 | 20 | 2007 |
| 10 | LIU Y | 10 | 14 | 0.588235 | 223 | 20 | 2007 |
| 11 | PRODGER JL | 8 | 13 | 0.666667 | 273 | 13 | 2012 |
| 12 | VELAZQUEZ EF | 13 | 14 | 0.619048 | 554 | 14 | 2003 |
| 13 | WANG H | 7 | 13 | 0.5 | 173 | 14 | 2010 |
| 14 | WANG J | 7 | 14 | 0.538462 | 221 | 16 | 2011 |
| 15 | WANG L | 6 | 14 | 0.352941 | 212 | 15 | 2007 |
| 16 | WANG X | 17 | 28 | 1 | 797 | 29 | 2007 |
| 17 | WANG Y | 11 | 18 | 0.647059 | 366 | 27 | 2007 |
| 18 | WU CS | 11 | 17 | 1.1 | 307 | 18 | 2014 |
| 19 | ZHANG Y | 14 | 20 | 0.933333 | 450 | 28 | 2009 |
| 20 | ZHOU J | 9 | 14 | 0.692308 | 233 | 14 | 2011 |

Supplement Table 2 The detailed of the top 10 citations from 2003 to 2022 in male prepuce

| Title | DOI | Year | GCS |
| --- | --- | --- | --- |
| A CULTURE SYSTEM USING HUMAN FORESKIN FIBROBLASTS AS FEEDER CELLS ALLOWS PRODUCTION OF HUMAN EMBRYONIC STEM CELLS | 10.1093/humrep/deg290 | 2003 | 365 |
| HUMAN FEEDER LAYERS FOR HUMAN EMBRYONIC STEM CELLS | 10.1095/biolreprod.102.012583 | 2003 | 334 |
| DERIVATION OF HUMAN EMBRYONIC STEM CELL LINES IN SERUM REPLACEMENT MEDIUM USING POSTNATAL HUMAN FIBROBLASTS AS FEEDER CELLS | 10.1634/stemcells.2004-0201 | 2005 | 185 |
| HIV-1 TARGET CELLS IN FORESKINS OF AFRICAN MEN WITH VARYING HISTORIES OF SEXUALLY TRANSMITTED INFECTIONS | 10.1309/JVHQVDJDYKM58EPH | 2006 | 81 |
| POTENTIAL HIV-1 TARGET CELLS IN THE HUMAN PENIS | 10.1097/01.aids.0000237364.11123.98 | 2006 | 169 |
| FINE-TOUCH PRESSURE THRESHOLDS IN THE ADULT PENIS | 10.1111/j.1464-410X.2006.06685.x | 2007 | 110 |
| FORESKIN SURFACE AREA AND HIV ACQUISITION IN RAKAI, UGANDA (SIZE MATTERS) | 10.1097/QAD.0b013e328330eda8 | 2009 | 74 |
| WITHIN 1 H, HIV-1 USES VIRAL SYNAPSES TO ENTER EFFICIENTLY THE INNER, BUT NOT OUTER, FORESKIN MUCOSA AND ENGAGES LANGERHANS-T CELL CONJUGATES | 10.1038/mi.2010.32 | 2010 | 108 |
| ABUNDANT EXPRESSION OF HIV TARGET CELLS AND C-TYPE LECTIN RECEPTORS IN THE FORESKIN TISSUE OF YOUNG KENYAN MEN | 10.2353/ajpath.2010.090926 | 2010 | 52 |
| MALE CIRCUMCISION | 10.1542/peds.2012-1990 | 2012 | 253 |
